# Subcortical brain volumes in neonatal hypoxic-ischemic encephalopathy

**DOI:** 10.1101/2022.12.06.22283178

**Authors:** Lilian M N Kebaya, Bhavya Kapoor, Paula Camila Mayorga, Paige Meyerink, Kathryn Foglton, Talal Altamimi, Emily S. Nichols, Sandrine de Ribaupierre, Soume Bhattacharya, Leandro Tristao, Michael T Jurkiewicz, Emma G. Duerden

## Abstract

**Background:** Hypoxic ischemic encephalopathy (HIE) is a severe brain injury impacting term-born neonates. Despite treatment with therapeutic hypothermia (TH), HIE is associated with myriad adverse developmental outcomes suggesting the involvement of subcortical structures, including the thalamus and basal ganglia, which may be vulnerable to perinatal asphyxia, particularly during the acute period.

**Aims:** 1) To examine subcortical macrostructure in the first few days of life in neonates with HIE compared to age- and sex-matched healthy neonates. 2) To determine whether subcortical volumetric maturation is associated with HIE severity.

**Methods:** A cohort of 28 neonates (19 males [67.9%], median gestational age [GA]=38.6 weeks, interquartile range [IQR]=36.8-39.6) with HIE (mild=4, moderate=21, severe=3 based on Sarnat Staging) were scanned with MRI within the first four days of life (median postmenstrual age [PMA]=39.2, IQR=37.6-40.3), with the majority of scans occurring in the post-cooling period (n=23[82%]). The control group included 28 healthy neonates matched for GA, birth weight and PMA at the scan. Subcortical volumes (thalamus, basal ganglia, hippocampus, cerebellum) were automatically extracted from T1-weighted images. General linear models assessed between-group differences in subcortical volumes, adjusting for sex, GA, PMA, and total cerebral volumes. Within-group analyses evaluated the association between subcortical volumes and HIE severity.

**Results:** Neonates with HIE had significantly smaller bilateral thalamic, basal ganglia and right hippocampal and cerebellar volumes compared to healthy neonates (all, p<0.02). Within the HIE group, milder HIE severity was associated with smaller volumes of the left and right basal ganglia (both, p<0.007) and the left hippocampus and thalamus (both, p<0.04) when adjusting for TH, days of mechanical ventilation and other clinical and demographic factors.

**Conclusions:** Consistent with findings from childhood survivors of HIE, newborns with HIE, scanned with MRI within the first days of life, had smaller subcortical volumes impacting sensory and motor regions, including the thalamus, basal ganglia and cerebellum compared to healthy newborns. Additionally, HIE severity was associated with subcortical volumes, particularly impacting the basal ganglia, suggesting these regions may be important brain-based biomarkers in newborns impacted by the hypoxic-ischaemic injury. Findings suggest that despite advances in neonatal care, HIE is associated with significant alterations in brain macrostructure.

## Introduction

Hypoxic-ischemic encephalopathy (HIE) due to perinatal asphyxia is a condition that causes a reduction of blood and oxygen to the neonatal brain and can lead to brain damage and adverse developmental outcomes, including cerebral palsy, cognitive impairment and epilepsy. (1-3) Adverse developmental outcomes seen in childhood survivors are often widespread, impacting multiple domains of function, indicating the involvement of subcortical structures with widespread connectivity to cortical areas.

HIE is highly prevalent and affects 1.5 per 1,000 live-born term neonates in developing countries. (4) The modified Sarnat examination is the most frequently used tool to assess the severity of HIE. (5, 6) Staging is based on evidence of the following criteria: history of a perinatal event, prolonged resuscitation, poor cord gases, low Apgar scores and findings consistent with encephalopathy, such as an abnormal neurological examination or seizures within the first 6 hours after birth. (5) HIE does not only cause immediate neuronal cell death (primary phase) but precipitates a complex biochemical cascade leading to delayed neuronal loss (secondary phase). (7) This conceptual framework forms the basis for the timely introduction of therapeutic hypothermia (TH, also known as cooling at 33.5°C ± 0.5°C) - to reduce further cell death and neuronal loss. TH started within the first 6 hours after birth, for 72 hours, is now the standard of care for moderate to severe HIE in developed countries. TH is currently the only treatment that has shown neuroprotective benefit. (8) To optimize neurodevelopmental outcomes, infants being treated with TH undergo intensive monitoring, including neuromonitoring (seizure detection and treatment, analgesia and sedation), cardio-respiratory monitoring (respiratory support if needed, blood pressure maintenance), nutrition and correction of metabolic derangements. (9) After 72 hours, infants are gradually rewarmed; after that, magnetic resonance imaging (MRI) is typically performed to characterize brain structure and injury.

In the acute period, hypoxic-ischaemic injury associated with HIE leads to characteristic patterns of injury impacting subcortical structures, including the hippocampus, thalamus, and basal ganglia identified on neonatal MRI scans. (10) A pilot study of volumetric MRI for participants with HIE found relatively larger volumes of subcortical white matter in infants treated with TH than in controls. (11) Shapiro et al., using deformation-based morphometry in infants with HIE at six months, showed local changes in the perisylvian grey and white matter that were associated with language outcomes at 30 months of age. (12) Children and adults treated for neonatal HIE also demonstrate altered hippocampal volumes. (13) Previous studies have also shown reduced total brain and cortical volumes in children with HIE compared with controls. (14) Presently, studies examining macrostructure volumes of infants with HIE in the acute period are scarce due to the challenges faced by their complex care. In turn, less is known about neonatal volumes in newborns with HIE; however, the acute period represents a critical window when brain macrostructure may be most vulnerable to hypoxic-ischaemic injury.

In the current study, based on previous research indicating that infants and children diagnosed with HIE at birth suffered long-term neurological consequences, we wished to examine brain development in neonates with HIE during the acute period. In a prospective cohort study, we examined the subcortical macrostructure (thalamus, basal ganglia, hippocampus, amygdala, cerebellum). Our first aim was to examine subcortical macrostructure in infants with HIE in the post-cooling period (within the first four days of life), compared to healthy term-born babies scanned at a comparable postmenstrual age (PMA), adjusting for demographic variables. The second aim of the study was to examine whether the severity of HIE, based on Sarnat staging, is associated with changes in subcortical macrostructure, adjusting for clinical factors related to systemic illness and demographic variables.

## Methods

### Use of human subjects

This study was conducted as part of an ongoing prospective study of infants at risk of neurological injury. All participants were recruited from the Neonatal Intensive Care Unit (NICU) at the Children’s Hospital of South Western Ontario, London, Canada. The study was approved by the Health Sciences Research Ethics Board at Western University. Informed consent was provided by the parents/caregivers of the infants enrolled in the study.

### Participants

Infants with HIE were eligible for recruitment in this prospective study between January 2020 and June 2022. The following criteria were required for inclusion in the sub-study: gestational age ≥ 36 weeks, with a birth weight≥ 2000 g and admitted with a diagnosis of HIE. (5) HIE diagnosis was based on the following criteria: cord gas pH of ≤7.0 and/or base deficit of ≥16 mmol per L; if pH was between 7.01 and 7.15 or a base deficit was between 10 and 15.9 mmol per L, additional history of an acute perinatal event and an APGAR score at 10 minutes of ≤5, or need for assisted ventilation/resuscitation at birth; the presence of seizures or moderate or severe encephalopathy by standardized neurological examination. (15) Infants were excluded if they had evidence of major anomalies of the brain or other organs, congenital infections (e.g., TORCH), intrauterine growth restriction (IUGR), identifiable metabolic disorder or ultrasound evidence of a large parenchymal haemorrhagic infarction.

### Controls

In addition to the participants with HIE, the study included 28 healthy newborns with no reported brain injury in the analysis. The participants were infants born at term from the publicly available Developing Human Brain Connectome Project (DHCP, Hughes et al. 2017). DHCP is the first open-access data release of brain images in healthy neonates born at term who had an MRI scan within the first few days of life (37–44 weeks of gestational age), comparable with the HIE participants. With these data, we had access to the T1-weighted images. Additional MRI data-acquisition-related information is included in Supplementary Information. The images from the healthy newborns were selected based on participants’ gestational age and age at the MRI scan to ensure minimal variability.

### Clinical and Demographic Variables

Maternal and infant data were abstracted from electronic medical records. Demographic data included gestational age, birth weight, sex, HIE stage, mode of delivery, place of birth, resuscitation details, Apgar scores and cord pH. Details of 72 h treatment with TH and the presence of brain injury on MRI were collected. Infants with HIE underwent at least one MRI scan during hospitalization (after rewarming). If infants underwent multiple scans, imaging obtained immediately after rewarming was prioritized for scoring.

Days on ventilation (invasive and non-invasive) were recorded. Invasive ventilation: a sum of the days on conventional, jet or high-frequency oscillatory ventilation. Non-invasive ventilation: continuous positive pressure ventilation (CPAP), high- or low-flow oxygen. Additional variables of interest included postnatal infections (clinical sepsis or positive culture infection, confirmed necrotizing enterocolitis) and brain injury identified on MRI (intraventricular haemorrhage [IVH], white-matter injury [WMI]).

### MRI Image Acquisition

All HIE participants underwent an MRI scan as part of clinical care on day 4 of life (median day of life [DOL] at scan=4, interquartile range [IQR]=4.4). All scans were obtained while participants were resting or asleep. None of the infants was sedated for the MRI scan. Structural MRI scans were acquired on a 1.5 T 450 W GE scanner. Each infant underwent a clinical MRI scan consisting at a minimum of a whole-brain T1-weighted structural image (TR=8.4–11.5 ms, depending on clinical requirements, TE=4.2 ms, flip angle=12/25°, matrix size 512 × 512, 99–268 slices, voxel size typically 0.39 × 0.39 × 0.5 mm (0.31 × 31 × 5 to 0.43 × 0.43 × 0.6 for some infants), and a T2-weighted structural image (TR=3517–9832 ms, TE=7.3–8.4 ms, flip angle = 90/160°, matrix size 256 × 256, 19–60 slices, 0.7 × 0.7 × 2–5 mm voxel resolution). Alternative imaging sequences such as EPI (resting-state functional MRI), FLAIR, and SWAN sequences were added if requested by the attending physician.

#### Brain injury characterization

A pediatric neuroradiology fellow (LT) scored the T1-weighted anatomical images for brain injury severity (see Figure 1). These were verified by a paediatric neuroradiologist (MJ). White matter injury (WMI) was defined as foci exhibiting T1 hyperintensity without T2 hypointensity or by low-intensity T1 foci and was scored on a 3-point scale (none=0, minimal=1, moderate-severe=2-3 combined) using the methods of de Vries (1992). (16) Intraventricular haemorrhage (IVH) was graded (none=0, mild=1-2, and moderate-severe=3-4) using Papile’s method. (17) Only supratentorial injuries were scored.

**Figure 1:**
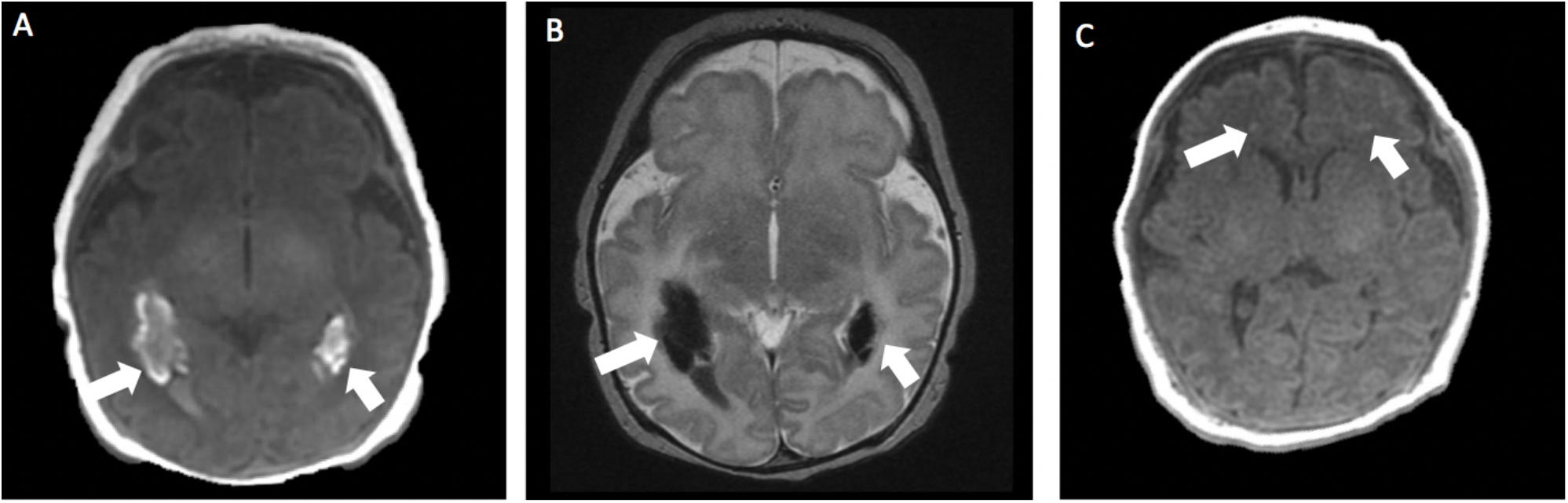
Brain injury patterns on T1 weighted images: A: Intraventricular haemorrhage in the trigone of the lateral ventricles as seen on a T1 weighted image. B: Intraventricular haemorrhage seen on a T2 weighted image. C: Punctate white matter injury as seen on a T1 weighted image.

### Processing and Segmentation

The raw DICOM data were converted to NifTI files through MRIcroGL. The resulting T1 weighted imaging scans converted to 3D images were segmented automatically using infant FreeSurfer. (18) Infant FreeSurfer is an automatic processing stream for T1-weighted MRI scans in infants. (18) Automatic processing steps include intensity normalization, skull stripping, and segmentation of the cortex, white matter and subcortical structures. (18) Intensity normalization involves standardizing MRI signal intensity which may differ across participants, scanners, or visits. (19) Skull-stripping is the process of removing all non-brain tissue from MRI images. Segmentation involves segmenting the brain into its anatomical features, including cortical/subcortical brain structures and white/grey matter.

The Freesurfer method involves a multi-atlas approach, in which multiple atlases of newborns are first registered to native space and structure labels are transferred. The atlases were developed from infant MRI scans. (20) To initially create the atlases, manually-segmented labels were developed using MRI scans from a representative sample of infants (0-2 years of age). In the current study, developmentally appropriate atlases for newborns were employed. Labels are fused into a single segmentation result, providing higher accuracy than single-atlas approaches. (21) Volumetric measurements for anatomical features can then be extracted. (18)

Infant brain subcortical brain structure volumes were extracted as well as cerebral white matter and cerebral cortex volumes, to compute total cerebral volumes (TCV). Subcortical brain structures included the left and right: cerebellum cortex, thalamus, caudate, putamen, pallidum, hippocampus, amygdala, accumbens area and ventral diencephalon. The ventricles (lateral, third, and fourth) were also automatically labelled. An example of automatic segmentation of a newborn with HIE and a healthy newborn with T1-weighted MRI scans is provided in Figure 2. Each segmented T1-weighted image was visually inspected by one of the authors (BK) using the Freeview software, available within the FreeSurfer suite of tools. Seven (25%) T1-weighted images underwent additional manual relabelling. Further manual segmentation was employed to correct any segmentation errors (i.e., partial volume effects) in the subcortical grey matter using ITK-SNAP (http://www.itksnap.org/).

**Figure 2:**
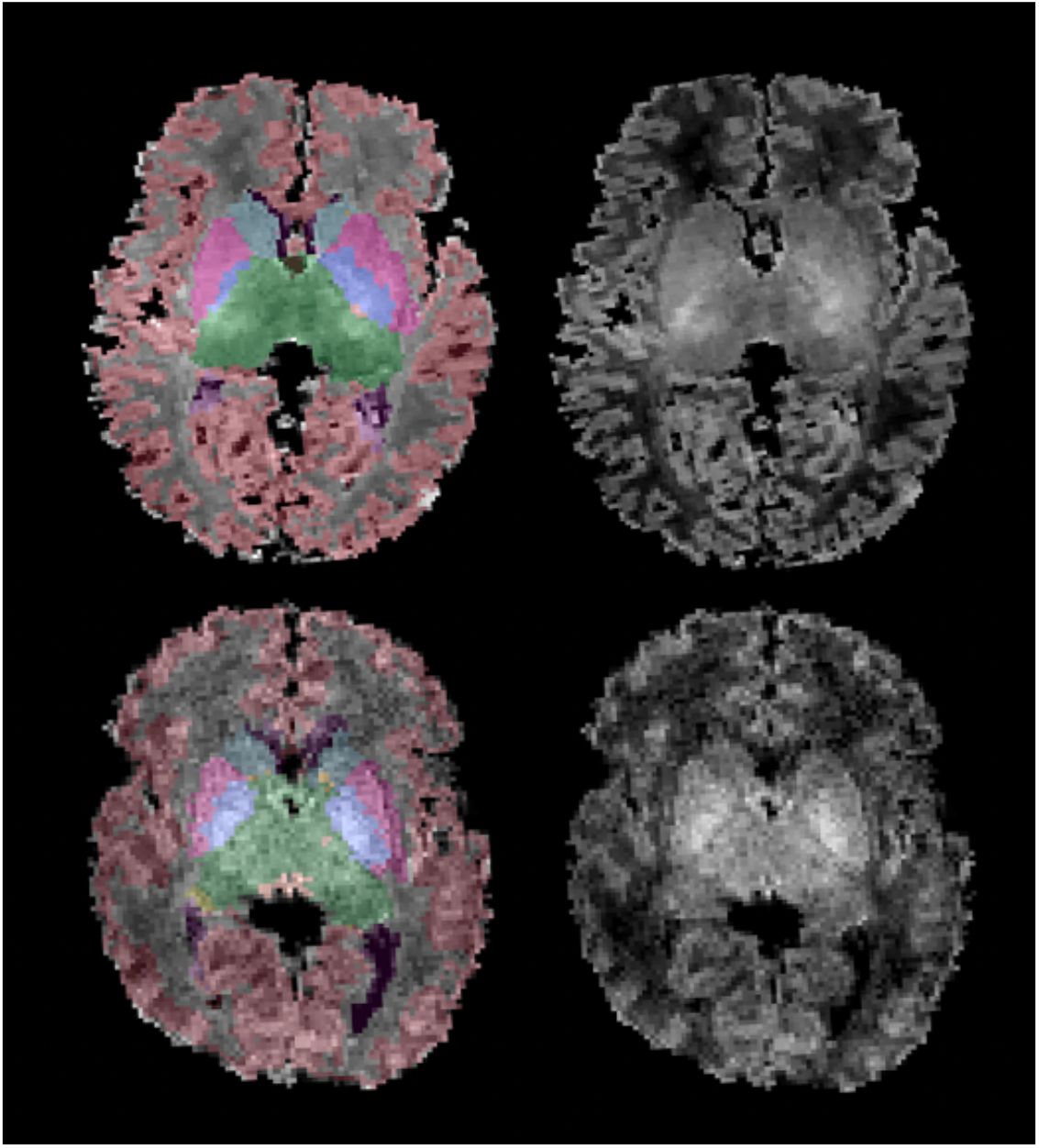
Segmented T-1 weighted images comparing HIE and control subjects. The top row of images is from a newborn with HIE. The second row is the control. Both subjects had no other brain injuries found.

### Statistical Analysis

All statistical analyses were performed using Statistical Package for the Social Sciences (v.27 [SPSS], Chicago, IL). The main aim of the study was to assess volumetric differences in subcortical structures between newborns with HIE compared to healthy newborns. Two multivariate models were run to address the first aim of the study. In the first model, the volumes of the left and right basal ganglia (summed values of the caudate, putamen, and pallidum), thalamus, hippocampus, amygdala, and cerebellum were the dependent variables and the independent variable was group (HIE, healthy newborn), adjusting for sex, gestational age, PMA at the scan and total cerebral volume. As a single was run to assess a single hypothesis, with subcortical volumes being smaller in the HIE group compared to healthy newborns, the alpha level was set at p=0.05.

The second aim of the study was to examine whether the severity of HIE was associated with subcortical development. A basic model including the dependent variables (left and right basal ganglia, thalamus, hippocampus, amygdala, cerebellum) and HIE severity was entered as a continuous variable to examine the heterogeneity of the disorder in relation to brain macrostructure, rather than making between-group comparisons. The model was adjusted for TH (yes/no), sex, gestational age, total cerebral volume and total number of days on mechanical ventilation. As we had a single hypothesis regarding the association of HIE severity and larger subcortical volumes, the alpha level was set at p=0.05.

## Results

### Participants

We identified 33 infants with HIE. Five were excluded due to unavailable MRI (early death or discharge home) or poor image quality resulting from significant motion artefacts. Hence, our study comprised 28 newborns (19 male, nine female) with HIE. Of the 28 participants, three were diagnosed with mild HIE, 21 with moderate HIE, and four with severe HIE in accordance with Sarnat staging. (3)

#### Therapeutic Hypothermia

Of the 28 infants, 23 underwent whole body TH treatment for 72 hours at 33.5°C, followed by gradual re-warming over 6 hours (at 0.5°C per hour) until the core temperature reached 36.5°C, as per unit policy.

#### Clinical characteristics

The HIE patient and healthy newborn groups were matched for sex, gestational age, PMA at scan and birth weight (Table 1). The HIE participants included in the study had a variety of clinical criteria assessed (Table 2). Twelve (42.9%) of the 28 newborns were outborn (born at another facility or home) and then transferred to our NICU. The mean Apgar at 5 minutes was 4.35 ± 1.05, and the mean first pH was 6.97 ± 0.05. Half of the study population required intubation at birth. We also recorded the total number of days on ventilation (invasive or non-invasive) as the mean number of days on invasive ventilation, non-invasive ventilation, or a combination of the 3 (4.54 ± 1.89). Most newborns had their scans on the fourth day following rewarming.

**Table 1:** Participant Characteristics

|  | **HIE participants**<br>**N=28** | **Healthy newborns**<br>**N=28** | **P value** |
| --- | --- | --- | --- |
| Gestational Age, *weeks [IQR]* | 38.5 [36.4- 39.8] | 38.5 [36.6-39.7] | 0.9 |
| Sex, *male n, (%)* | 19 (67.9) | 19 (67.9) | 0.9 |
| Birth Weight, *kgs [IQR]* | 3.38 [2.53-3.79] | 3.15 [2.58-3.61] | 0.3 |
| PMA at scan, *weeks [IQR]* | 38.9 [36.98-40.2] | 38.8 [37.5-40.3] | 0.9 |

The median values and interquartile ranges. Probability values provide results using the t-test for continuous measures and Chi-square tests for categorical measures. PMA: postmenstrual age

**Table 2:** Descriptive and clinical data

|  | **HIE newborns (*n* = 28)** | **P value** |
| --- | --- | --- |
| Place of birth | Inborn: 16 (57.1%)<br>Outborn: 12 (42.9%) | 0.45 |
| Delivery method | Spontaneous vaginal delivery: 17 (60.7%)<br>Caesarean section: 11 (39.3%) | 0.257 |
| First pH<br>(cord or peripheral) | Mean = $6.97 \pm 0.05$,<br>Min = 6.8, Max = 7.3, SD = 0.14<br>25<sup>th</sup> Percentile: up to 6.9<br>50<sup>th</sup> percentile: up to 6.915<br>75<sup>th</sup> percentile: up to 7.0375 | |
| Apgar 5 min | Mean = $4.35 \pm 1.05$,<br>Min = 0, Max = 9, SD = 2.74<br>25<sup>th</sup> Percentile: up to 2<br>50<sup>th</sup> percentile: up to 5<br>75<sup>th</sup> percentile: up to 6 | |
| Sarnat staging | Mild: 3 (10.7%)<br>Moderate: 21 (75.0%)<br>Severe: 4 (14.3%) | <0.0001 |
| Therapeutic<br>hypothermia | Yes: 23 (82.1%)<br>No: 5 (17.9%) | 0.001 |
| Intubation at birth | Yes: 14 (50.0%)<br>No: 14 (50.0%) | 0.9 |
| Days of ventilation | Mean = $4.54 \pm 1.89$ days,<br>Min = 0 days, Max = 21 days, SD = 5.11 days<br>25<sup>th</sup> Percentile: up to 1.0 days<br>50<sup>th</sup> percentile: up to 1.5 days<br>75<sup>th</sup> percentile: up to 8.0 days | |

#### Brain Injury

Of the 28 infants with HIE, 27 (96%) had at least mild injury on MRI at a median age of 5 (IQR 4–6) days. Grade 1 (mild) white matter injury was commonly seen in the neonates, regardless of HIE severity (n=22, 79%). Basal ganglia injury was evident in two (7%) neonates with severe HIE, while IVH was seen in two (7%) participants, bilateral grade IV in one (3.6%) participant with mild HIE, and bilateral grade II in one participant (3.6%) with moderate HIE. More than one injury pattern was seen in four (14%) neonates. No evidence of watershed injury was evident in any of the neonates with HIE (Table 3).

**Table 3:**
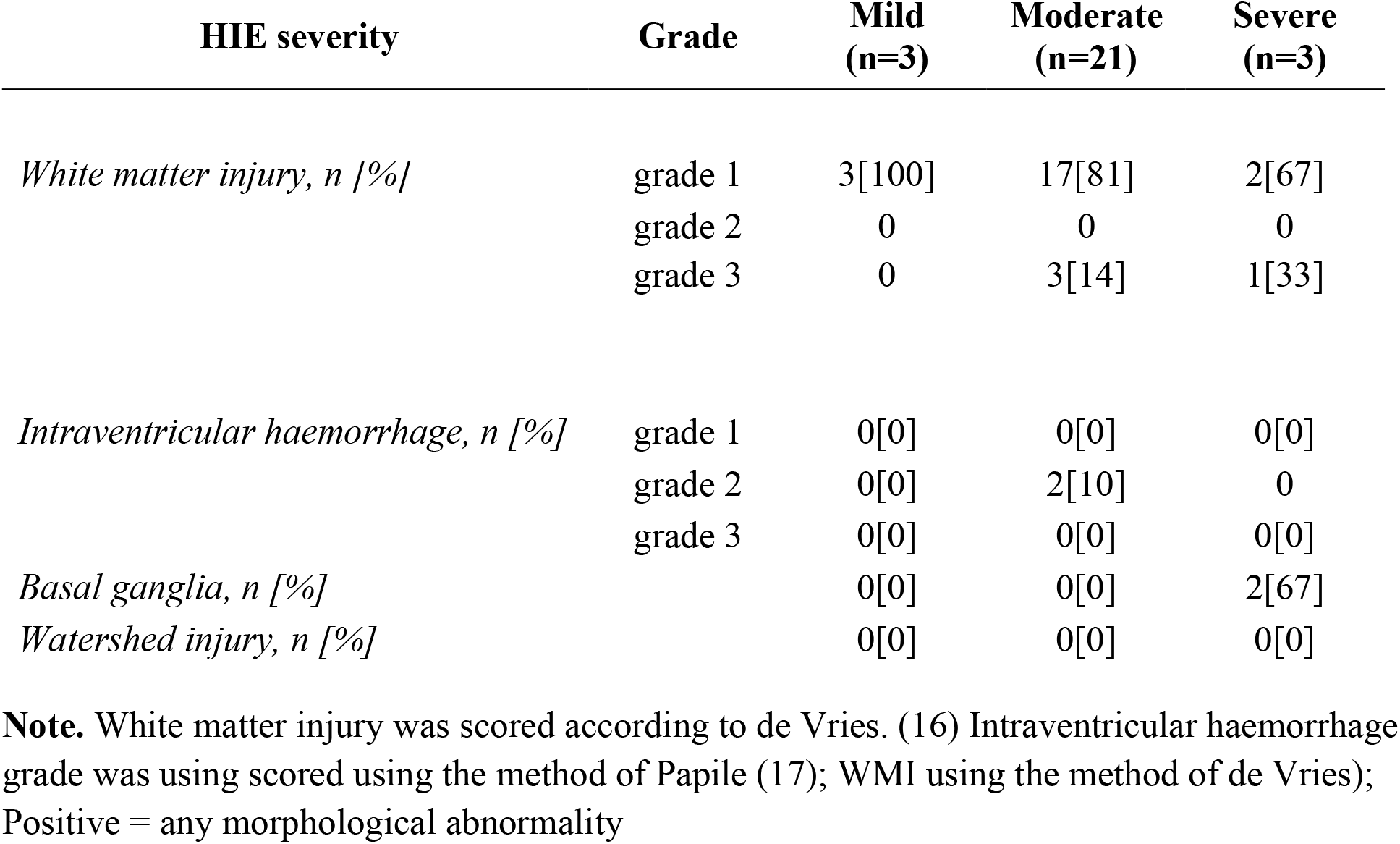
Brain injury patterns in HIE participants

| HIE severity | Grade | Mild<br>(n=3) | Moderate<br>(n=21) | Severe<br>(n=3) |
| --- | --- | --- | --- | --- |
| *White matter injury, n [%]* | grade 1 | 3[100] | 17[81] | 2[67] |
|  | grade 2 | 0 | 0 | 0 |
|  | grade 3 | 0 | 3[14] | 1[33] |
| *Intraventricular haemorrhage, n [%]* | grade 1 | 0[0] | 0[0] | 0[0] |
|  | grade 2 | 0[0] | 2[10] | 0 |
|  | grade 3 | 0[0] | 0[0] | 0[0] |
| *Basal ganglia, n [%]* |  | 0[0] | 0[0] | 2[67] |
| *Watershed injury, n [%]* |  | 0[0] | 0[0] | 0[0] |

**Note.** White matter injury was scored according to de Vries. (16) Intraventricular haemorrhage grade was using scored using the method of Papile (17); WMI using the method of de Vries); Positive = any morphological abnormality

### Between groups analysis: Relationship between subcortical macrostructure and HIE diagnosis

A multivariate model revealed a main effect of the group (F=[6,42]9.84, p<0.001). Newborns with HIE had significantly smaller right (F=16.2, df=1, p<0.001) and left thalamic (F=10.8, df=1, p=0.002) volumes compared to typically-developing newborns (Fig. 3, Table 4). The right (F=34.4, df=1, p<0.001) and left basal ganglia (F=12.6, df=1, p<0.001) as well as the right hippocampus (F=41.3, df=1, p<0.001) and right cerebellar (F=5.7, df=1, p=0.02) volumes were also smaller in the newborns with HIE compared to healthy newborns. Volumetric differences in amygdala volumes were not evident (p>0.05).

**Figure 3:**
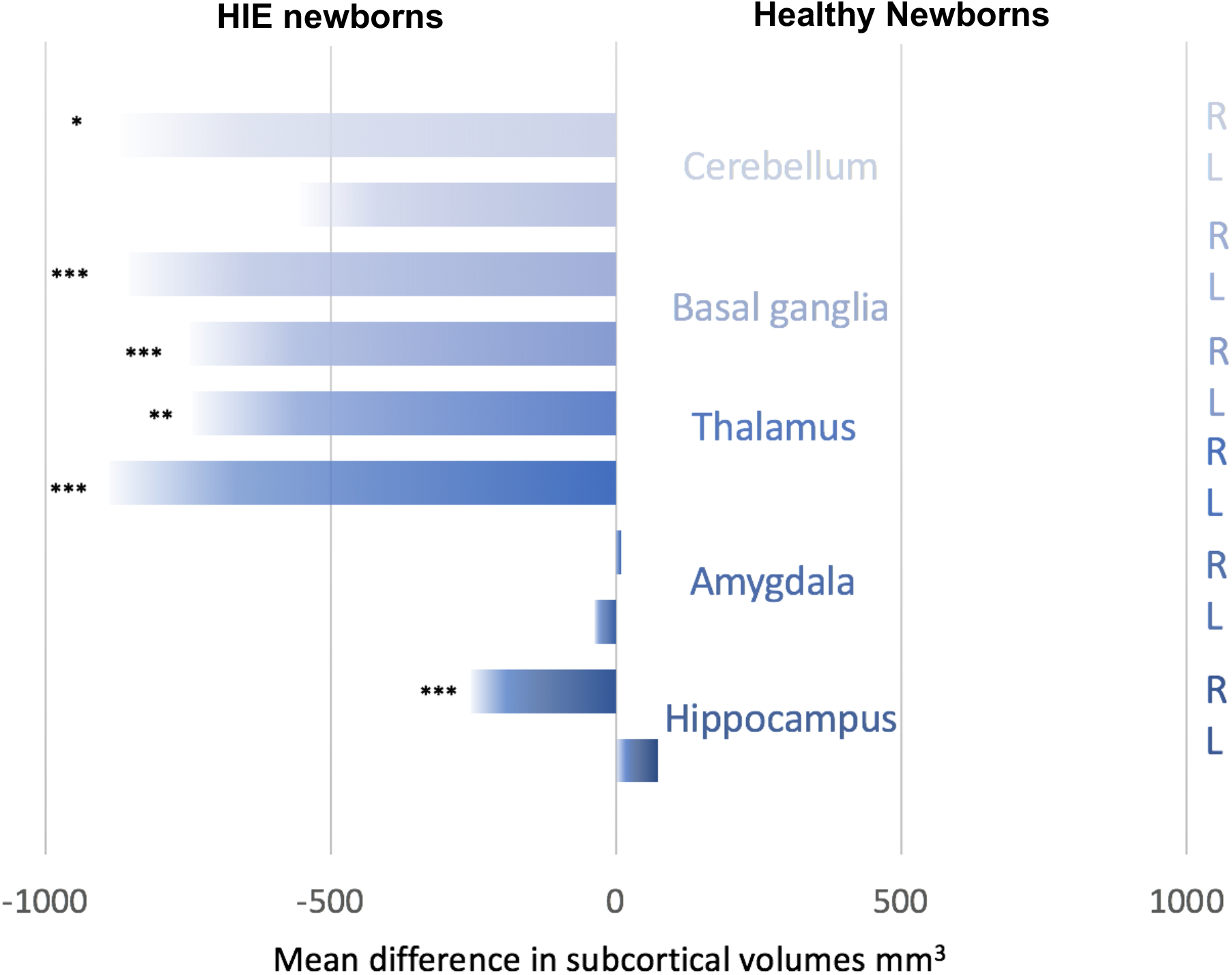
Subcortical macrostructural differences in mean volumes between participants with HIE compared to healthy newborns. Values represent the output from a general linear model and are the mean differences in volumes between the HIE group and healthy newborns, adjusting for birth gestational age, age at scan, sex and total cerebral volumes. *** p<0.001, **p<0.01, *p<0.05

**Table 4:** Results of a multivariate general linear model comparing subcortical volumes between HIE newborns and controls

| Brain Volume | Side | df | F value | P value |
| --- | --- | --- | --- | --- |
| Basal Ganglia | **Right** | **1** | **34.3** | **<.001** |
|  | **Left** | **1** | **12.6** | **<.001** |
| Hippocampus | **Right** | **1** | **41.3** | **<.001** |
|  | Left | 1 | 3.0 | 0.09 |
| Amygdala | Right | 1 | 0.1 | 0.8 |
|  | Left | 1 | 2.0 | 0.2 |
| Thalamus | **Right** | **1** | **10.8** | **0.002** |
|  | **Left** | **1** | **16.2** | **<.001** |
| Cerebellum | **Right** | **1** | **5.7** | **0.02** |
|  | Left | 1 | 1.9 | 0.2 |

Results of a multivariate general linear model comparing brain volumes between the HIE and healthy newborn group adjusting for sex, birth gestational age, PMA at the scan and total cerebral volumes. Significant values are in bold.

### Within-group analysis: Relationship between subcortical macrostructure and HIE severity

To address our second aim, subcortical volumes were examined in relation to HIE severity based on Sarnat staging while adjusting for sex, gestational age, TH and days of mechanical ventilation, and total cerebral volumes. A multivariate model revealed a main effect of HIE severity (F= [10,7]4.34, p=0.032). The left (F=9.68, p=0.007) and right (F=15.11, p=0.001) basal ganglia, the left hippocampus (F=5.78, p=0.029), and the left thalamus (F=4.8, p=0.044) were associated with HIE severity (Table 5). Posthoc univariate general linear models revealed that HIE severity was associated with increased volumes in all structures, including the left (B=1409.3, df=1, p=0.008) and right (B=1268.2, df=1, p<0.001) basal ganglia, left hippocampus (B=291.3, df=1, p=0.008), and the left (B=1084.9, df=1, p=0.047) thalamus, adjusting for the same clinical and demographic covariates.

**Table 5:** Results of a multivariate general linear model examining subcortical volumes in relation to HIE severity

| Brain Volume | Side | df | F value | P value |
| --- | --- | --- | --- | --- |
| Basal ganglia | Left | 1 | **9.68** | **0.007** |
|  | Right | 1 | **15.11** | **0.001** |
| Hippocampus | Left | 1 | **5.78** | **0.03** |
|  | Right | 1 | 1.71 | 0.21 |
| Amygdala | Left | 1 | 3.59 | 0.08 |
|  | Right | 1 | 2.59 | 0.13 |
| Thalamus | Left | 1 | **4.80** | **0.04** |
|  | Right | 1 | 4.6 | 0.05 |
| Cerebellum | Left | 1 | 0.33 | 0.57 |
|  | Right | 1 | 0.69 | 0.42 |

Results of a multivariate general linear model examining subcortical brain volumes in HIE newborns in relation to severity based on Sarnat staging. The model was adjusted for sex, birth gestational age, TH, days of mechanical ventilation and total cerebral volumes. Significant values are in bold.

## Discussion

Hypoxic ischemic encephalopathy remains prevalent and is associated with myriad adverse neurodevelopmental outcomes. Despite advances in neonatal care, including therapeutic hypothermia, HIE is associated with significant alterations in brain macrostructure. We compared the subcortical brain macrostructure between neonates with HIE and their healthy counterparts. The initial volumetric comparisons demonstrated reductions in the basal ganglia, thalami, and right hippocampal and cerebellar volumes in patients with HIE. Further, we found that HIE severity was associated with basal ganglia hippocampal and thalamic volumes. Assessment of brain injury in the acute period is vital to determine the severity of the disease and prognosis. Hence, subcortical brain areas could be important brain-based markers for the early identification of neonates at risk for long-term neurodevelopmental impairment. Moreover, the findings of this study bear implications, especially since an infant’s brain experiences rapid brain growth during the first year of life, during which the brain is most amenable to early therapeutic interventions. (22)

Volumetric analysis showed reduced volumes in the basal ganglia, thalami, right cerebellum and right hippocampus in the HIE group. Our findings are aligned with previous studies. A prospective study with preterm infants (with and without perinatal HIE) found decreased cerebellum and brainstem volumes at term-equivalent age compared to healthy term-born infants, a finding that persisted at two years. (23) Although the HIE population was preterm (GA 23-32 weeks), a similar inclusion criterion was used in addition to neurological deficits and abnormal radiological findings that were consistent with HIE. Additionally, none of the babies received TH. The application of TH in preterm neonates (< 36 weeks GA) remains unclear and is currently being studied. (24) Nonetheless, their findings support the results of our study. Spencer et al. reported reduced thalamic, hippocampal, and grey matter volumes in children aged 6-8 (31 patients and 32 controls) treated with TH for neonatal HIE. Although their study was done outside infancy, it further shows that subcortical brain volume loss in HIE survivors persists even at school-going age and is associated with decreased functional outcomes. (25) Other childhood studies have consistently reported similar findings. (13, 14)

Our findings of reduced basal ganglia and thalami (BGT) volumes support well-known animal and clinical evidence showing BGT injury as one of the predominant patterns of hypoxic-ischemic injury. (2, 10, 26, 27) Deep grey matter regions, including basal ganglia and thalamus, are particularly vulnerable to hypoxic-ischemic injury due to their high metabolic rate, ongoing myelination and high concentration of glutamate receptors, making them prone to volume loss following hypoxic-ischemic injury. (26, 28) Adult studies following traumatic brain injury show subcortical volume loss in the thalamus and basal ganglia, further reinforcing that these regions are likely to suffer volume loss. (29) Information regarding decreased BGT volumes is important, given the importance of these regions in sensorimotor functions. The thalamus is the conduit of all sensory and motor information from the peripheral nervous system to the cerebral cortex. (30) Previous studies have consistently reported that neonates with reduced basal ganglia volumes experience more developmental delays and deficits than their healthy counterparts. (31-35) Smaller subcortical structures are associated with poor motor and higher-order executive functions, as found in the 7-year follow-up of neonates born with HIE. (36)

We also found that HIE severity was associated with subcortical volumes, particularly impacting the basal ganglia, adjusting for clinical and demographic factors, including TH. Existing animal evidence shows that increasing hypoxic-ischemic injury severity is associated with a shorter latent phase, worse secondary energy failure and more severe cortical damage. (37) Further, existing human evidence demonstrates more severe acute neurological injury, even death, with greater HIE severity, TH notwithstanding. (37, 38) The basal ganglia are particularly vulnerable to hypoxic-ischemic injury. The reason is that glutamate receptors are prevalent in the basal ganglia, and glutamate-induced toxicity plays an important role in ischemic injury in term infants, which can ultimately result in neuronal cell death. Second, the neonatal brain, particularly the BGT, is sensitive to oxidative injury following perinatal asphyxia. All these factors, combined with mitochondrial dysfunction and the release of inflammatory processes, contribute to cellular alterations, including synaptic pruning that could impact subcortical macrostructure. Microglia play an important role in controlling synaptic pruning in a healthy developing brain. In HIE, however, this process could be impaired, leading to inappropriate synaptic connections. (39) Besides the known association of HIE targeting the basal ganglia resulting in overt motor deficits (40), there is also an increased risk for neuropsychiatric disorders in the long term. Basal ganglia injury is associated with up to 7-fold increased risk of developing schizophrenia in HIE survivors. (42) Neuropsychiatric disorders are associated with larger subcortical volumes; (41) however, the association between BGT injury and later development of neuropsychiatric disorders remains an active area of investigation. Improved information concerning subcortical volumetric development within the first week of life in newborns impacted by HIE provides an opportunity to identify neonates who will benefit from close interdisciplinary follow-up or additional neuroimaging.

In the current study, all newborns had MRI scans completed within the first week of life. We included a comparable control group in terms of baseline characteristics. This permitted the investigation of subcortical macrostructure in infants with HIE in the post-cooling period. Although our study offers new insights into brain subcortical volumes in newborns with HIE, the findings should be considered with respect to some limitations. First, with a relatively small sample size of 28 newborns with HIE, the conclusions drawn require more research to support them. However, HIE at our centre is seen in relatively low incidence, and to recruit newborns for an extensive period, during which time changes in clinical care may occur, could unduly influence results in relation to changes in brain macrostructure. Future prospective studies with larger sample sizes are needed. Lastly, our study only reports imaging findings in the acute period with no linking to neurodevelopmental outcomes.

## Conclusions

Consistent with findings from childhood survivors of HIE, newborns with HIE, scanned with MRI within the first days of life, had smaller subcortical volumes impacting sensory and motor regions, including the thalamus, basal ganglia and cerebellum, compared to healthy newborns. Additionally, HIE severity was associated with larger subcortical volumes, particularly impacting the basal ganglia, suggesting these regions may be important brain-based biomarkers in newborns impacted by the hypoxic-ischaemic injury. These findings suggest that despite advances in neonatal care, HIE is associated with significant alterations in brain macrostructure.

## Data Availability

The families who participated were not consented to share the data publically.

## Authors contributions

LMNK, BK, PCM, PM, KF, TA, ESN, SB, SdR, LT, MJ, and EGD were involved in the study design, database variable creation, and data acquisition design and execution of the data analytic strategy, reviewed and/or revised the final version of the manuscript.

LMNK, BK, and EGD contributed to the execution of the data analytic strategy, analyzed the data and wrote the initial draft of the manuscript.

All authors approved the final manuscript as submitted and agree to be accountable for all aspects of the work.

## Acknowledgements

We thank the NICU families who participated in this study.

## Data Availability Statement

The data supporting this study’s findings are available from the corresponding author upon reasonable request.

## Supplementary Information

### DHCP MRI Protocol

The data from the DHCP cohort were collected at the Evelina Neonatal Imaging Center, London, on a 3T Philips Achieva scanner using a 32-channel phased array dedicated neonatal head coil. An inversion recovery T1-weighted multi-slice fast spin-echo images was acquired in sagittal and axial slice stacks with in-plane resolution 0.8mm by 0.8mm and 1.6mm thick slices overlapped by 0.8mm. T1 weighted sequence was acquired with a TR of 4795ms, inversion time of 1740ms and TE of 8.7ms along with axial SENSE factor of 2.27 and sagittal SENSE factor of 2.66

## References

1. Lee AC, Kozuki N, Blencowe H, Vos T, Bahalim A, Darmstadt GL, et al. Intrapartum-related neonatal encephalopathy incidence and impairment at regional and global levels for 2010 with trends from 1990. Pediatr Res. 2013;74 Suppl 1:50–72.

2. Gunn AJ, Bennet L. Fetal hypoxia insults and patterns of brain injury: insights from animal models. Clin Perinatol. 2009;36(3):579–93.

3. Lee BL, Glass HC. Cognitive outcomes in late childhood and adolescence of neonatal hypoxic-ischemic encephalopathy. Clin Exp Pediatr. 2021;64(12):608–18.

4. Kurinczuk JJ, White-Koning M, Badawi N. Epidemiology of neonatal encephalopathy and hypoxic-ischaemic encephalopathy. Early Hum Dev. 2010;86(6):329–38.

5. Sarnat HB, Sarnat MS. Neonatal encephalopathy following fetal distress. A clinical and electroencephalographic study. Arch Neurol. 1976;33(10):696–705.

6. Mrelashvili A, Russ JB, Ferriero DM, Wusthoff CJ. The Sarnat score for neonatal encephalopathy: looking back and moving forward. Pediatr Res. 2020;88(6):824–5.

7. de Vries LS, Cowan FM. Evolving understanding of hypoxic-ischemic encephalopathy in the term infant. Semin Pediatr Neurol. 2009;16(4):216–25.

8. Jacobs SE, Berg M, Hunt R, Tarnow-Mordi WO, Inder TE, Davis PG. Cooling for newborns with hypoxic ischaemic encephalopathy. Cochrane Database Syst Rev. 2013(1): CD003311.

9. Wintermark P, Mohammad K, Bonifacio SL, Newborn Brain Society G, Publications Committee. Electronic address pno. Proposing a care practice bundle for neonatal encephalopathy during therapeutic hypothermia. Semin Fetal Neonatal Med. 2021;26(5):101303.

10. de Vries LS, Groenendaal F. Patterns of neonatal hypoxic-ischaemic brain injury. Neuroradiology. 2010;52(6):555–66.

11. Parikh NA, Lasky RE, Garza CN, Bonfante-Mejia E, Shankaran S, Tyson JE. Volumetric and anatomical MRI for hypoxic-ischemic encephalopathy: relationship to hypothermia therapy and neurosensory impairments. J Perinatol. 2009;29(2):143–9.

12. Shapiro KA, Kim H, Mandelli ML, Rogers EE, Gano D, Ferriero DM, et al. Early changes in brain structure correlate with language outcomes in children with neonatal encephalopathy. Neuroimage Clin. 2017;15:572–80.

13. Annink KV, de Vries LS, Groenendaal F, van den Heuvel MP, van Haren NEM, Swaab H, et al. The long-term effect of perinatal asphyxia on hippocampal volumes. Pediatr Res. 2019;85(1):43–9.

14. Bregant T, Rados M, Vasung L, Derganc M, Evans AC, Neubauer D, et al. Region-specific reduction in brain volume in young adults with perinatal hypoxic-ischaemic encephalopathy. Eur J Paediatr Neurol. 2013;17(6):608–14.

15. Lemyre B, Chau V. Hypothermia for newborns with hypoxic-ischemic encephalopathy. Paediatr Child Health. 2018;23(4):285–91.

16. de Vries LS, Eken P, Dubowitz LM. The spectrum of leukomalacia using cranial ultrasound. Behav Brain Res. 1992;49(1):1–6.

17. Papile LA, Burstein J, Burstein R, Koffler H. Incidence and evolution of subependymal and intraventricular haemorrhage: a study of infants with birth weights less than 1,500 gm. J Pediatr. 1978;92(4):529–34.

18. Zollei L, Iglesias JE, Ou Y, Grant PE, Fischl B. Infant FreeSurfer: An automated segmentation and surface extraction pipeline for T1-weighted neuroimaging data of infants 0-2 years. Neuroimage. 2020;218:116946.

19. Sun X, Shi L, Luo Y, Yang W, Li H, Liang P, et al. Histogram-based normalization technique on human brain magnetic resonance images from different acquisitions. Biomed Eng Online. 2015;14:73.

20. de Macedo Rodrigues K, Ben-Avi E, Sliva DD, Choe MS, Drottar M, Wang R, et al. A FreeSurfer-compliant consistent manual segmentation of infant brains spanning the 0-2 year age range. Front Hum Neurosci. 2015;9:21.

21. Iglesias JE, Sabuncu MR. Multi-atlas segmentation of biomedical images: A survey. Med Image Anal. 2015;24(1):205–19.

22. Knickmeyer RC, Gouttard S, Kang C, Evans D, Wilber K, Smith JK, et al. A structural MRI study of human brain development from birth to 2 years. J Neurosci. 2008;28(47):12176–82.

23. Raguz M, Rados M, Kostovic Srzetic M, Kovacic N, Zunic Isasegi I, Benjak V, et al. Structural Changes in the Cortico-Ponto-Cerebellar Axis at Birth are Associated with Abnormal Neurological Outcomes in Childhood. Clin Neuroradiol. 2021;31(4):1005–20.

24. Sabir H, Bonifacio SL, Gunn AJ, Thoresen M, Chalak LF, Newborn Brain Society G, et al. Unanswered questions regarding therapeutic hypothermia for neonates with neonatal encephalopathy. Semin Fetal Neonatal Med. 2021;26(5):101257.

25. Spencer APC, Lee-Kelland R, Brooks JCW, Jary S, Tonks J, Cowan FM, et al. Brain volumes and functional outcomes in children without cerebral palsy after therapeutic hypothermia for neonatal hypoxic-ischaemic encephalopathy. Dev Med Child Neurol. 2022.

26. Miller SP, Ramaswamy V, Michelson D, Barkovich AJ, Holshouser B, Wycliffe N, et al. Patterns of brain injury in term neonatal encephalopathy. J Pediatr. 2005;146(4):453–60.

27. Misser SK, Lotz JW, Zaharie SD, McHunu N, Archary M, Barkovich AJ. A proposed magnetic resonance imaging grading system for the spectrum of central neonatal parasagittal hypoxic-ischaemic brain injury. Insights Imaging. 2022;13(1):11.

28. Roland EH, Poskitt K, Rodriguez E, Lupton BA, Hill A. Perinatal hypoxic-ischemic thalamic injury: clinical features and neuroimaging. Ann Neurol. 1998;44(2):161–6.

29. Gooijers J, Chalavi S, Beeckmans K, Michiels K, Lafosse C, Sunaert S, et al. Subcortical Volume Loss in the Thalamus, Putamen, and Pallidum, Induced by Traumatic Brain Injury, Is Associated With Motor Performance Deficits. Neurorehabil Neural Repair. 2016;30(7):603–14.

30. Torrico TJ, Munakomi S. Neuroanatomy, Thalamus. StatPearls. Treasure Island (FL)2022.

31. Schneider J, Duerden EG, Guo T, Ng K, Hagmann P, Bickle Graz M, et al. Procedural pain and oral glucose in preterm neonates: brain development and sex-specific effects. Pain. 2018;159(3):515–25.

32. Martinez-Biarge M, Diez-Sebastian J, Kapellou O, Gindner D, Allsop JM, Rutherford MA, et al. Predicting motor outcome and death in term hypoxic-ischemic encephalopathy. Neurology. 2011;76(24):2055–61.

33. Ilves N, Loo S, Ilves N, Laugesaar R, Loorits D, Kool P, et al. Ipsilesional volume loss of basal ganglia and thalamus is associated with poor hand function after ischemic perinatal stroke. BMC Neurol. 2022;22(1):23.

34. Nivins S, Kennedy E, Thompson B, Gamble GD, Alsweiler JM, Metcalfe R, et al. Associations between neonatal hypoglycaemia and brain volumes, cortical thickness and white matter microstructure in mid-childhood: An MRI study. Neuroimage Clin. 2022;33:102943.

35. Geva S, Jentschke S, Argyropoulos GPD, Chong WK, Gadian DG, Vargha-Khadem F. Volume reduction of caudate nucleus is associated with movement coordination deficits in patients with hippocampal atrophy due to perinatal hypoxia-ischaemia. Neuroimage Clin. 2020;28:102429.

36. Loh WY, Anderson PJ, Cheong JLY, Spittle AJ, Chen J, Lee KJ, et al. Neonatal basal ganglia and thalamic volumes: very preterm birth and 7-year neurodevelopmental outcomes. Pediatr Res. 2017;82(6):970–8.

37. Iwata O, Iwata S, Thornton JS, De Vita E, Bainbridge A, Herbert L, et al. “Therapeutic time window” duration decreases with increasing severity of cerebral hypoxia-ischaemia under normothermia and delayed hypothermia in newborn piglets. Brain Res. 2007;1154:173–80.

38. Morales MM, Montaldo P, Ivain P, Pant S, Kumar V, Krishnan V, et al. Association of Total Sarnat Score with brain injury and neurodevelopmental outcomes after neonatal encephalopathy. Arch Dis Child Fetal Neonatal Ed. 2021;106(6):669–72.

39. Paolicelli RC, Bolasco G, Pagani F, Maggi L, Scianni M, Panzanelli P, et al. Synaptic pruning by microglia is necessary for normal brain development. Science. 2011;333(6048):1456–8.

40. Perlman JM. Intrapartum hypoxic-ischemic cerebral injury and subsequent cerebral palsy: medicolegal issues. Pediatrics. 1997;99(6):851–9.

41. Hagberg H, Gressens P, Mallard C. Inflammation during fetal and neonatal life: implications for neurologic and neuropsychiatric disease in children and adults. Ann Neurol. 2012;71(4):444–57.

42. Ursini G, Punzi G, Chen Q, Marenco S, Robinson JF, Porcelli A, et al. Convergence of placenta biology and genetic risk for schizophrenia. Nat Med. 2018;24(6):792–801.

